# Studying the progress of COVID-19 outbreak in India using SIRD model

**DOI:** 10.1101/2020.05.11.20098681

**Authors:** Saptarshi Chatterjee, Apurba Sarkar, Swarnajit Chatterjee, Mintu Karmakar, Raja Paul

## Abstract

We explore a standard epidemiological model, known as the SIRD model, to study the COVID-19 infection in India, and a few other countries around the world. We use (a) the stable cumulative infection of various countries and (b) the number of infection versus the tests carried out to evaluate the model. The time-dependent infection rate is set in the model to obtain the best fit with the available data. The model is simulated aiming to project the probable features of the infection in India, various Indian states, and other countries. India imposed an early lockdown to contain the infection that can be treated by its healthcare system. We find that with the current infection rate and containment measures, the total active infection in India would be maximum at the end of June or beginning of July 2020. With proper containment measures in the infected zones and social distancing, the infection is expected to fall considerably from August. If the containment measures are relaxed before the arrival of the peak infection, more people from the susceptible population will fall sick as the infection is expected to see a three-fold rise at the peak. If the relaxation is given a month after the peak infection, a second peak with a moderate infection will follow. However, a gradual relaxation of the lockdown started well ahead of the peak infection, leads to a nearly two-fold increase of the peak infection with no second peak. The model is further extended to incorporate the infection arising from the population showing no symptoms. The preliminary finding suggests that random testing needs to be carried out within the asymptomatic population to contain the spread of the disease. Our model provides a semi-quantitative overview of the progression of COVID-19 in India, with model projections reasonably replicating the current progress. The projection of the model is highly sensitive to the choice of the parameters and the available data. Besides, since the pandemic is an ongoing dynamic phenomenon, the reported results are subjected to regular updates in consonance with the acquired real data.

## INTRODUCTION

In the post-WW-2 era, the world probably has not witnessed such catastrophic morbidity and the looming threat of severe economic challenges caused by the worldwide outbreak of the disease COVID-19 caused by severe acute respiratory syndrome coronavirus 2 (SARS-CoV-2). The detection of the disease in the human host was first reported in Wuhan, China on 31 December 2019 as a cluster of cases of pneumonia. As the highly contagious disease transmitted rapidly all over the globe, the outbreak was declared as a pandemic by the WHO on 11 March 2020. Tackling the outspread of the disease is found to be very challenging across the world for the following reasons: (a) conventional flu-like symptoms in human carriers and (b) human-to-human transmission via asymptomatic human hosts and (c) the absence of a proper clinical doctrine (e.g. vaccines, drugs, concrete ideas about the immunological response, etc.). Extensive testing and the imposition of containment measures to maintain social distancing turn out to be the effective remedies to prevent disease transmission at the current stage of the epidemic at several places. To evaluate the impact of these preventive measures on infection spread, recovery, death tolls, and various other associated factors, mathematical models become useful in predicting realistic, quantitative estimates. A preliminary analysis suggests that the classic mean-field Susceptible-Infected-Recovery-Death (SIRD) model by Kermack and McKendrick [1, 2], can be used to obtain a quantitative picture of the epidemic [3–8]. In this article, implementing the SIRD model, we report the temporal progress of COVID-19 transmission in India, various Indian states and compare it with some other countries around the world. A similar model used by Fernandez-Villaverde et al [5] provides a detailed overview of the pandemic situation in the USA and many other countries.

India implemented a nation-wide lockdown from March 25, 2020. On the day of the announcement of nation-wide lockdown, India had about 657 corona positive cases, while the first COVID-19 positive was detected on January 30, 2020. The socio-economic constraints in the Indian context alludes that: (a) ‘too-prolonged’ lockdown is difficult to sustain; (b) the sole imposition of containment measures without a manifold increase in testing capacity is a futile endeavor; (c) if the implementation of the lockdown measures is lenient, containment of the spread is highly improbable. Henceforth, the feasible solution for limiting the spread lies in carefully balancing various key epidemiological factors. That is where the importance of the current model predictions becomes relevant.

This study further highlights the effect of lockdown on the disease spread and predictions about the variability in the infection peak upon the severity of the containment measures (and/or the lack of it). The model prediets that, in India, the height of the peak infection decreases with stricter lockdown, but at the cost of ‘time’ (position of the peak shifts to a later month). Thus, with a large susceptible, the infection will stay for a long time if existing infections are not quarantined immediately or no proper medicine/vaccine is employed. The key is to quarantine the infection in small pockets while in lockdown and prevent inter-pocket transmission. The model further underlines that in the highly contagious zones (‘red’ zones where COVID-19 positive cases continue to grow), if the lockdown is extended and enforced with proper quarantine measures, the new infections will gradually plummet down flattening the COVID-19 curve at a much faster rate.

Our study also explores the plausibility of universality in the spread of the COVID-19 outbreak amongst different countries [6] and compares the situation in India with few other countries (e.g. Germany, South Korea, USA, Spain) in the relevant time window (February - April). Due to the simplicity of the SIRD model, we found it difficult to fit the observed patterns of the pandemic using the available data. The real data for analysis in India’s context is collected from the repository with an interactive interface hosted at https://www.covid19india.org. The data for other countries are taken from the repository with an interactive interface hosted at https://www.worldometers.info/coronavirus. The purpose of this article is not to make any quantitative prediction that should be used to design policies, but for the research purpose only.

## MODEL

We employ the standard SIRD model where the population *N* is divided into sub-population of susceptible (S), infected (I), recovered (R) and dead (D) for all times *t*. Thus, *N* = *S+I* + *R* + *D*. The following set of mean-field differential equations governs the temporal dynamics of the population of susceptible (S), infected (I), recovered (R), dead (D) and describes a comprehensive picture of the SIRD epidemic evolution:

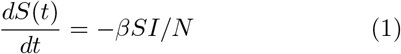

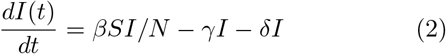

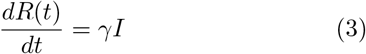

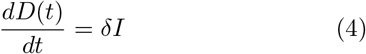

Here, *β*, *γ*, *δ* are the parameters determining the characteristics of infection, recovery and deaths respectively (Fig. 1A). Note that, in the current scenario *I* represents the population of symptomatic infection. When a susceptible person interacts with an infectious person, the susceptible become infected at a rate *βSI/N*. Large variability is observed in the rate *γ* that an infected individual is no longer infectious or equivalently has recovered in this simplified model. Literature [9–11] suggests that, on the average, infectiousness appears to start from 2-3 days before the symptoms are visible. The infectiousness increases to its peak before the arrival of the symptoms and remains for about 7-9 days after the peak infection. Thus an infected individual remains infectious for about 12 days on the average and then recover. In our preliminary analysis, we set the recovery rate *γ* ~ 1/12, which however does not give the best fit for all the cases we studied. In essence, the numerical values of the model parameters are obtained from the best fit. Initial values (time *t* = 0 days) of the number of infected, recovered and deaths (*I*_0_, *R*_0_*, D*_0_), are chosen from real data.

**FIG. 1.**
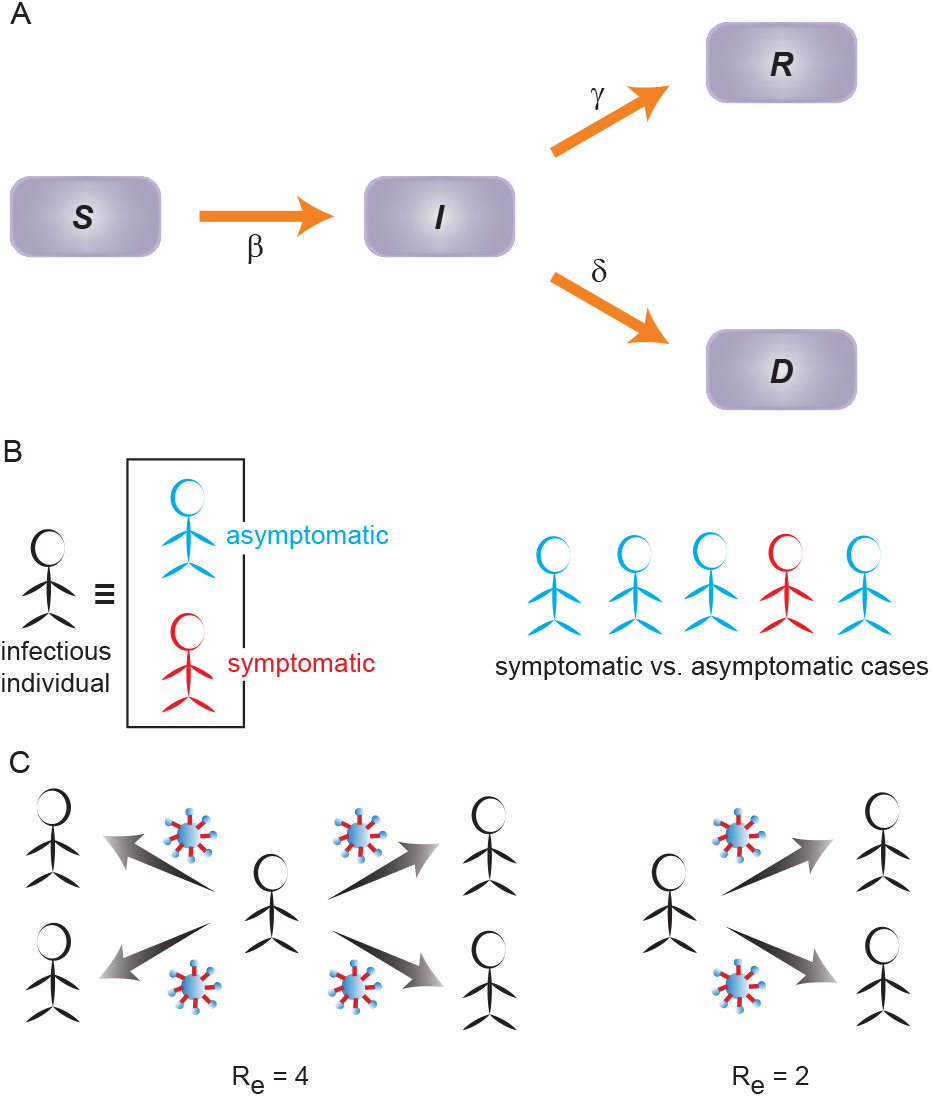
The transmission schematic representing the SIRD model. (A) The arrows indicate the flow between the populations of susceptible (S), infected (I), recovered (R), dead (D). (B) Infected individuals can be classified into two categories: symptomatic and asymptomatic. As per the WHO and the Indian Council of Medical Research (ICMR), India, the asymptomatic cases appears to be about 80% compared to the 20% that are symptomatic cases. (C) Effective reproduction number 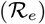: The average number of new infections transmitted by a single infectious person. For example, 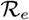 means one person is infecting 4 others on the average. Smaller the value of 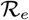, lesser the transmission capacity of the infection.

The choice for the initial number of susceptible (*S*_0_) is quite difficult. In the absence of antibody, the entire population can be susceptible to the COVID-19 pandemic. Nevertheless, the geographical, social, and economic characteristics of a region (and various other demographic factors) can substantially influence this number. We used two different approaches to get an estimate of *S*_0_. First, we study the data for the large countries where the cumulative positive cases have reached closer to a plateau. Though, the infected population at the plateau can be determined only when the epidemic is over. Dividing this number by the total population of the country gives a fraction that appears to be of the order for 10^−3^ for Germany, USA, Spain, Italy, and 10^−4^ and 10^−5^ for South Korea and China respectively. Thus an estimate of the susceptible may be obtained by multiplying the population of a country by this fraction. The number of susceptible obtained in this way, however, indicates a lower bound as many individuals with mild or no symptoms go unreported. Another possibility to estimate the fraction would be to test the number of positive cases by the number of tests carried out. This number would be an upper bound since there are many regions within a country that remains completely isolated and the populations in such pockets would not be susceptible. The ratio between the number of positive cases and the total number of tests for different countries are given in the following; the fraction is 0.159 for the USA, 0.016 for South Korea (as per data up to May 7), 0.1072 for Spain, 0.063 for Germany (as per data up to May 10). Conventionally, in epidemiological modeling *S*_0_ ~ *N*. In our simulation, we have reasonably varied *S*_0_ within this range to obtain the best fit with real data in a case by case manner (i.e for India, few Indian states and other countries).

With the formulation of the model, comes the quantitative estimate of the speed at which the disease spreads across a population. In other words, from the deterministic SIRD model, the objective is to assess how fast a human carrier would infect people belonging to the population of susceptible. The quantity that determines the transmission speed of the pandemic is the effective reproduction number or replacement number 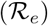[12]. Often the basic reproduction number 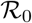, defined as the average number of secondary infections that occur when an infectious person (primary or source of infection) is placed into a susceptible population, is used in the epidemiological models. 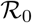 can be estimated from the very early stage of the infection when the infectious person mixes freely with the susceptible population. Estimating is often challenging due to lack of unbiased data as all secondary infections cannot be determined exactly; especially for COVID-19, where asymptomatic cases are hardly identified (Fig. 1B). The effective reproduction number 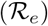, which we used in this study, evaluates the mean number of new infections (infected from the susceptible pool) directly transmitted/induced by a typical infected person and can vary over the entire duration of the infection (Fig. 1C). In the SIRD model, 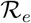 can be represented as *β*/(*γ*+*δ*). From the best fit of the data, we find that *γ >> δ*, yielding 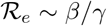. If 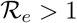, the disease starts spreading in a population infecting more and more people, but spreading does not occur if 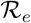 falls below 1. It is easy to notice that longer a person remains infectious (i.e. 1/*γ* days), can give rise to very large 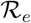 even if the number of infectious interactions per day (i.e. *β*) is small.

### Incorporating the effect of containment-measures

Containment measures in terms of social distancing and lockdown have been implemented world-wide to mitigate the transmission speed of the outbreak. We implemented the effect of lockdown in the model by modifying the infection rate and obtained the best-fit. We chose the following functional form of time-dependent infection rate 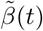 where it gradually decreases after the containment measures are enforced [5, 6]. Before lockdown, the infection rate is *β* which is constant. When the lockdown is imposed on day *τ* (counted from the initial time point *t* = 0 or day 0 as chosen in the simulation), the time-dependent infection rate 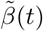 diminishes with every progressing day which is assume to vary exponentially in the following manner [6]:

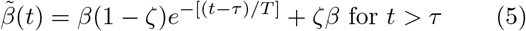

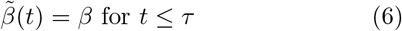

Here, ζ ∈ [0,1] is the infection parameter (or interaction parameter) and *T* is the delay in the number of days before the effect of lockdown is visible in the propagation of infection. Without lockdown ζ=1, referring to rapid infection while ζ = 0 means that infection is contained (e.g no interaction between infected and susceptible population, hence no transmission). ζ ∈ [0,1] reflects the asymptotic mitigation of the infection rate 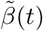, when containment measures are imposed. Lower the value of ζ, stricter is the containment measures (or the manifestation of the same) [5, 6]. Here the initially chosen value of *β* ends with ζ*β*. Essentially, the initial value of *β* determines the characteristic properties of the disease which depends on the effective interaction of people in a region, social behavior, density of population, etc. The terminal value ζ*β* reflects the effect of the containment and how the social distancing is being maintained. The functional form of 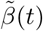 is arbitrary and chosen semi-empirically to obtain the best fit with the available data.

The model simulation, data analysis, and plotting are carried out in python. The analysis of the COVID-19 data, using the deterministic compartmental SIRD model, sheds light on the primary characteristics of the temporal evolution of the pandemic. Relevant parameter values chosen for the India and few Indian states are listed in Table S1-S2. The best-fit parameters chosen for foreign countries are listed in Table S3.

## RESULTS

We carried out the SIRD model analysis on COVID-19 progression in India’s context (and few other countries) with realistic variations in following parameters: rates of infection (*β*), recovery (*γ*) and deaths (*δ*), the initial number of susceptible (*S*_0_) and the effective reproduction number 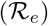. Detailed results are described in the following and illustrated in Fig. 2–10, Fig. S1-S2. In a nutshell, we start with the initial susceptible population (*S*_0_) varied within the range ~ 1-3 million, keeping the effective reproduction number 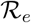 fixed at ~ 4.0, and show how the model prediction fits with the Indian data without a lockdown, the location of the infection peak and the relative deviation from the real data (Fig. 2A). The best fit is obtained by tuning the rates of infection (*β*), recovery (*γ*), and deaths (*δ*) keeping *S*_0_ constrained in the mentioned range. Then, we incorporate the effect of containment-measures/lockdown in the functional form of time-dependent 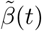 and show how the effect of the containment-measures has altered the location and the height of the infection peak (Fig. 2B). Next, we explore how the variability in the effective reproduction number 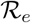 influences the infection peak (Fig. 3-4). Furthermore, we analyze the COVID-19 progression in few Indian states e.g Kerala, Maharashtra, Delhi, and West Bengal (Fig. 5-6) and foreign countries e.g South Korea, Germany, USA, Spain (Fig. 7-8). Lastly, we explore, in brief, what happens to the outspread, if the lockdown is lifted (in other words, containment measures are relaxed) in the Indian context (Fig. 9-10).

**FIG. 2.**
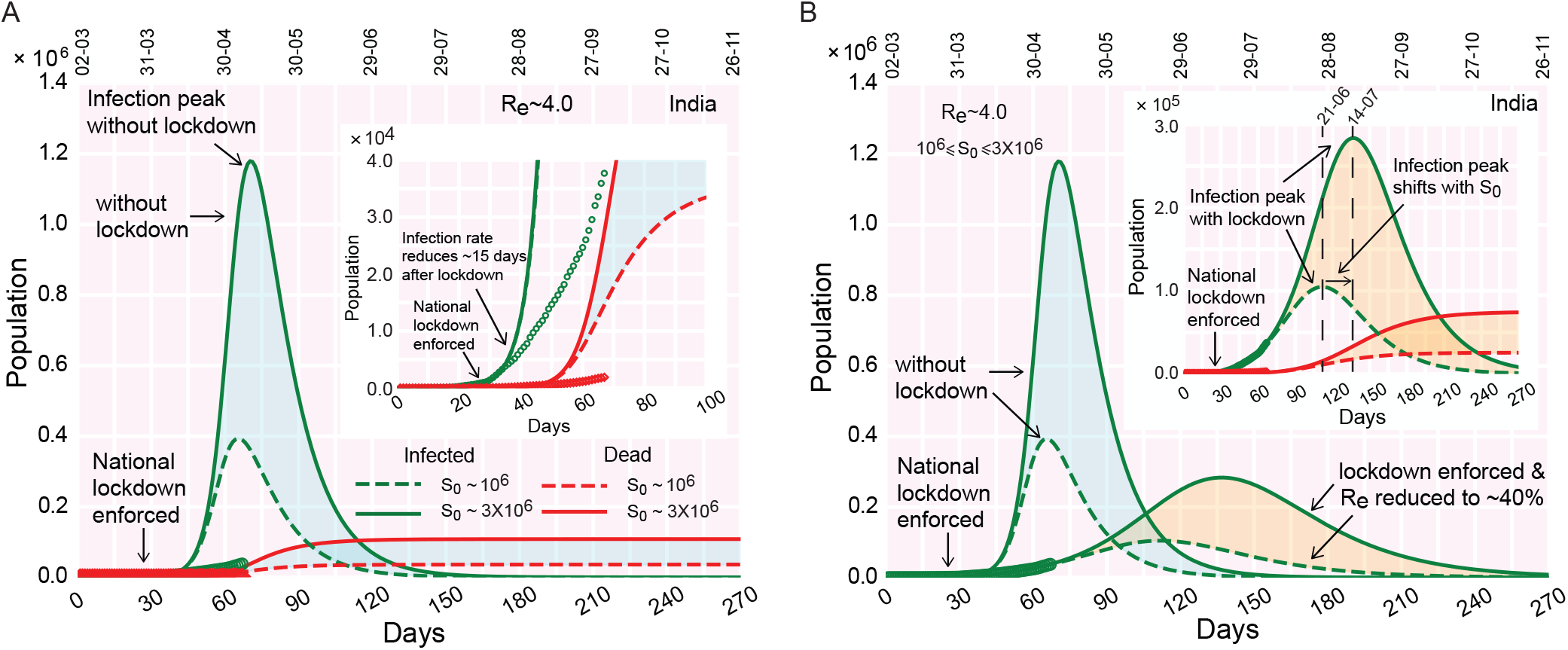
The infection growth curve and lockdown in India’s context. (A) Time progression of the infection growth curve, if no lockdown is imposed. (inset) The infection growth (as observed from the real data, points) reduces 20 days after the national lockdown is implemented deviating from the theoretical curve with no lockdown (dashed line). (B) Time evolution of the infected population (and deaths, inset), when containment measures (lockdown) are enforced. The color shades enveloping the curves denote the variation in susceptible population *S*_0_ (‘cyan’ shades: without lockdown, ‘orange’ shades: with lockdown). The real data considered for fitting are from March 2, 2020.

**FIG. 3.**
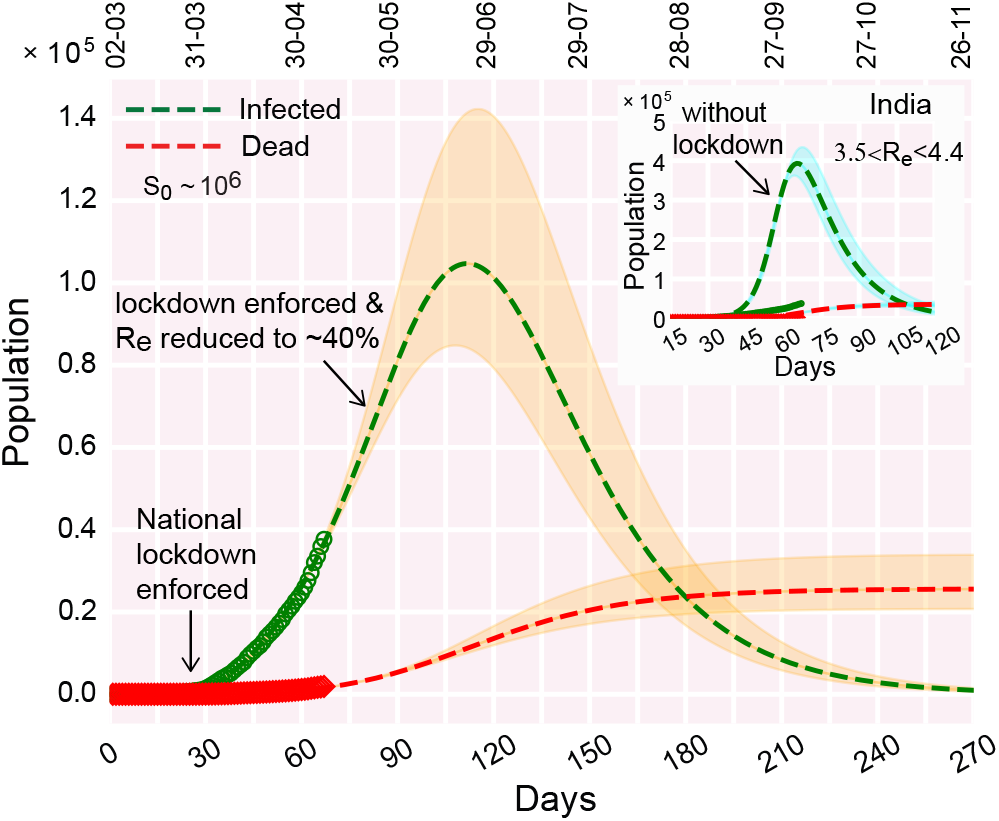
Effect of variations in 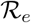 on the infection curve (and death curve).(A) Time progression of the infection growth curve when lockdown is enforced; (inset) if no lockdown was enforced. The color shades enveloping the curves denote the variation in 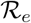. The real data considered for fitting are from March 2, 2020.

**FIG. 4.**
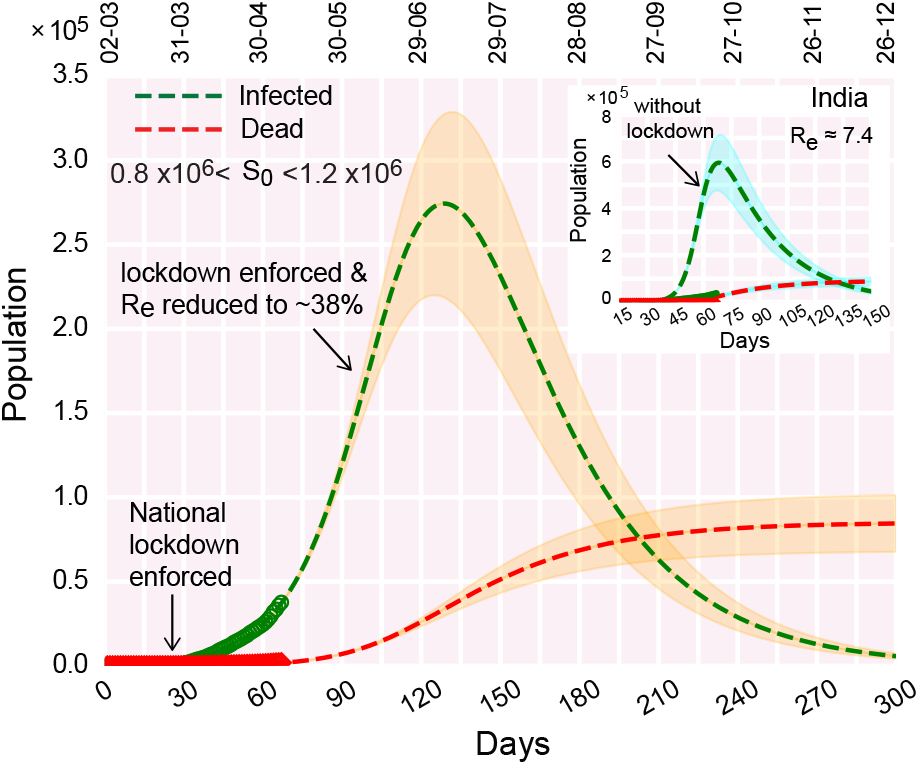
Best fit of the infection and death curves with the real data freely varying the effective 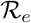 which appears to be ~ 7.4 in the early stage. The color shades enveloping the curves denote variation in susceptible population within a range of ~ 0.8 − 1.2 × 10^6^. The real data considered for fitting are from March 2, 2020. Relevant parameters for the analysis are *β* = 0.257 day^−1^, *γ* = 0.0315 day^−1^, *δ* = 0.0032 day^−1^, *τ* = 29 days, *T* = 7 days, ζ = 0.375.

**FIG. 5.**
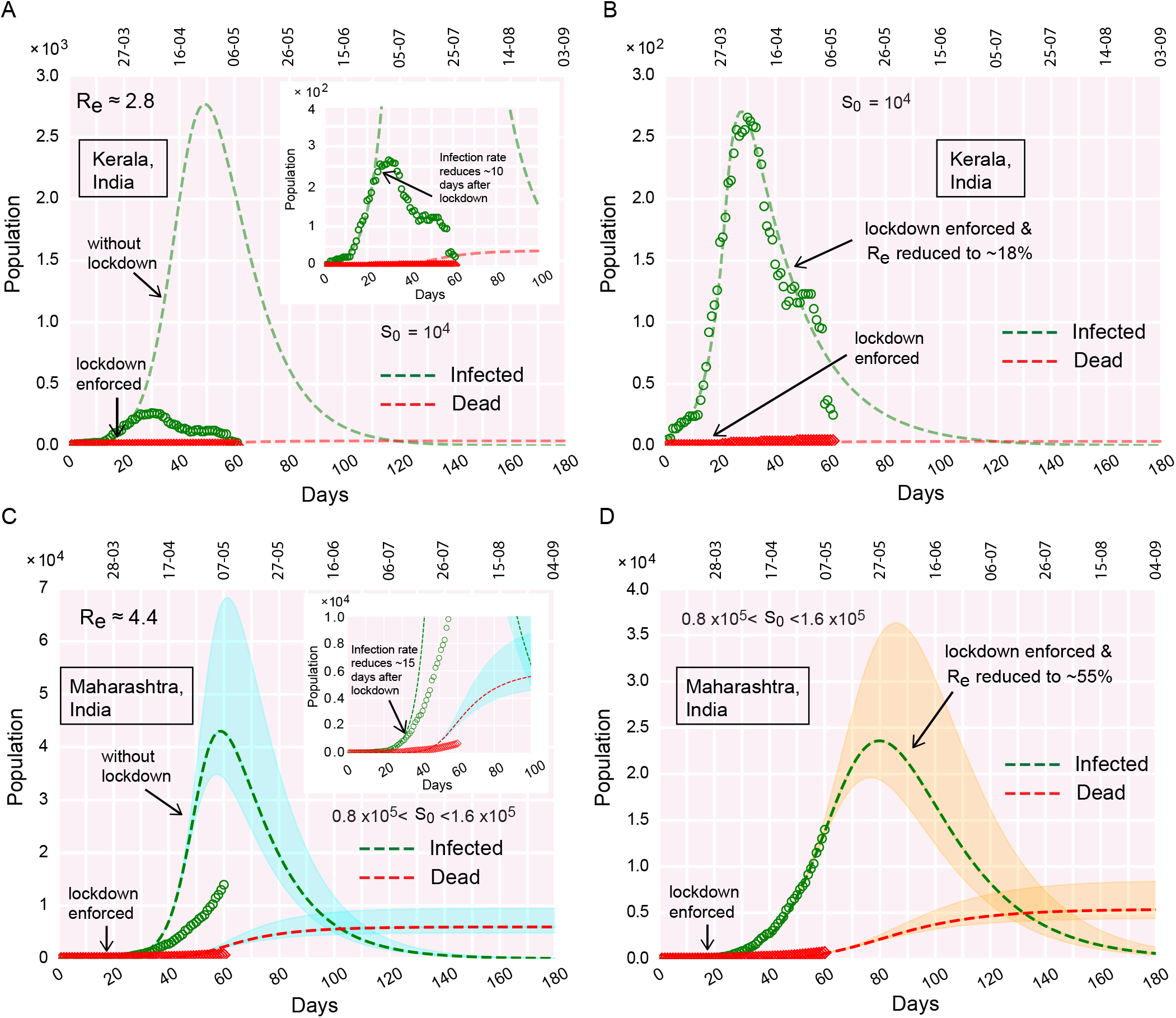
COVID-19 progression in few Indian states. (A-B) Time evolution of the population of infected and dead for Kerala, (A) if lockdown was not imposed, (B) due to effect of lockdown. (C-D) Time evolution of the population of infected and dead for Maharashtra, (C) if lockdown was not imposed, (D) due to effect of lockdown.

**FIG. 6.**
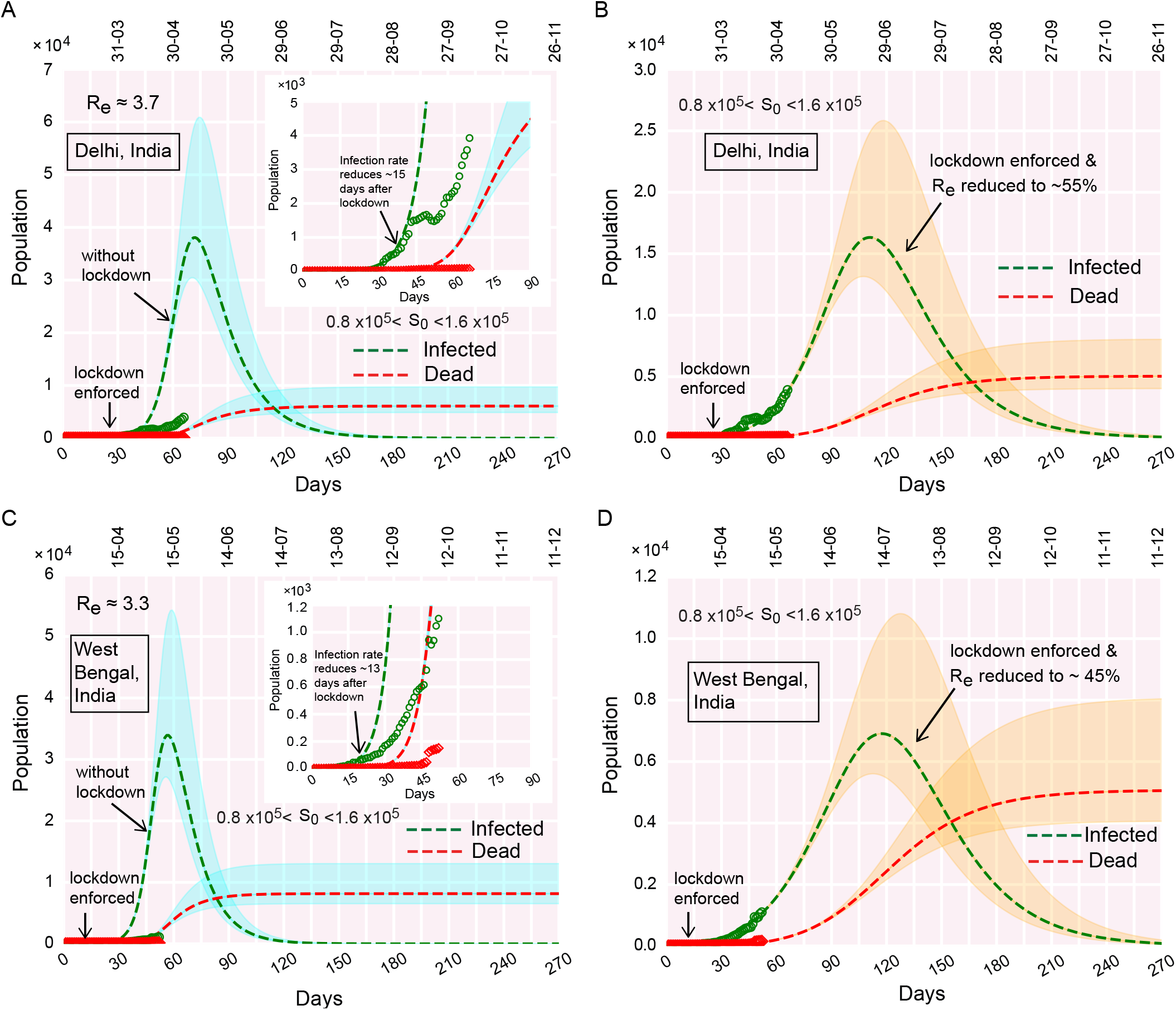
COVID-19 progression in few Indian states. (A-B) Time evolution of the population of infected and dead for Delhi, (A) if lockdown was not imposed, (B) due to effect of lockdown. (C-D) Time evolution of the population of infected and dead for West Bengal, (C) if lockdown was not imposed, (D) due to effect of lockdown.

**FIG. 7.**
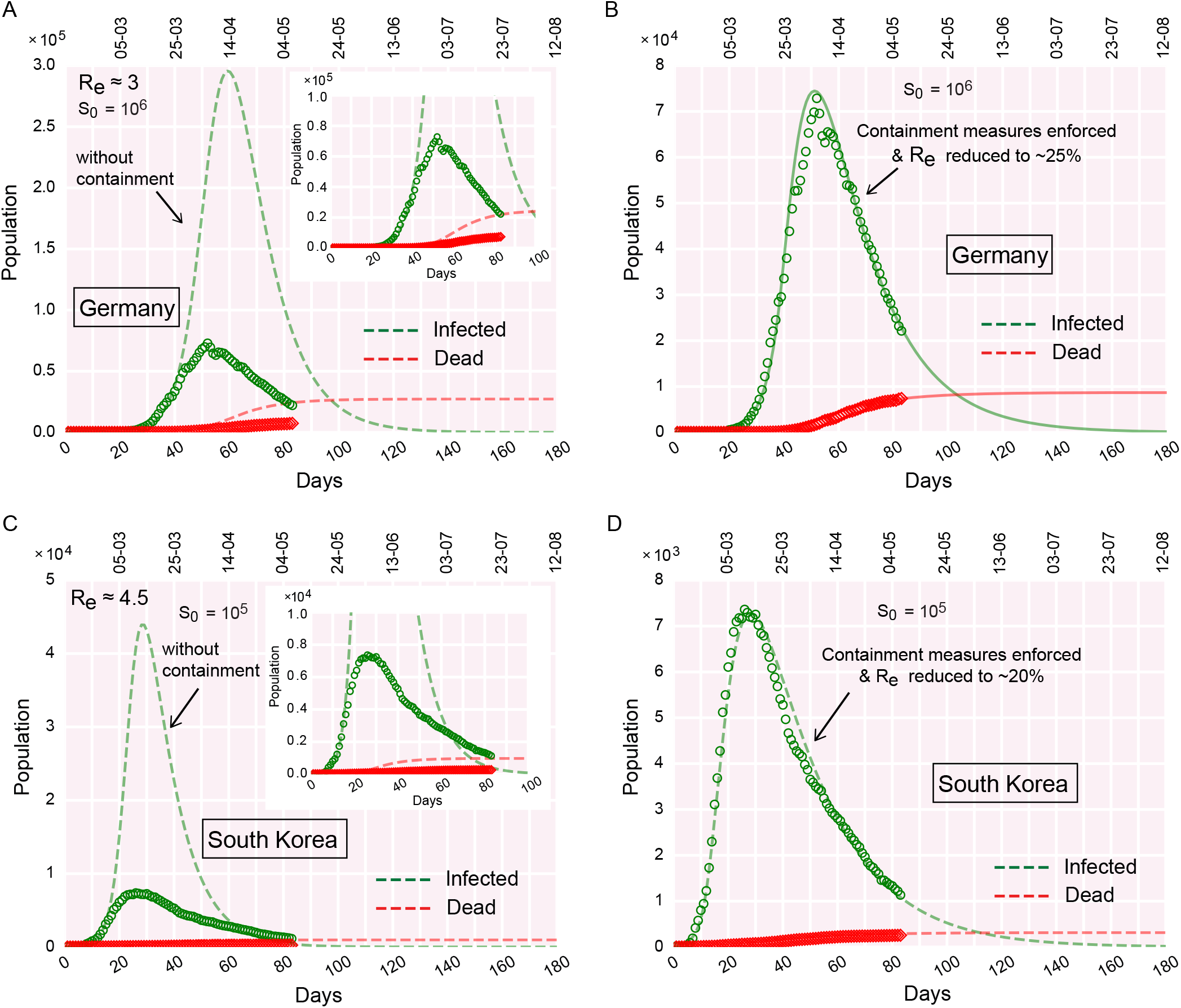
COVID-19 progression in Germany and South Korea. (A-B) Time evolution of the population of infected and dead for Germany, (A) if containment measures were not undertaken, (B) due to the effect of containment measures implemented. (C-D) Time evolution of the population of infected and dead for South Korea, (C) if containment measures were not undertaken, (D) due to the effect of containment measures implemented. The real data considered for fitting are from February 15, 2020.

**FIG. 8.**
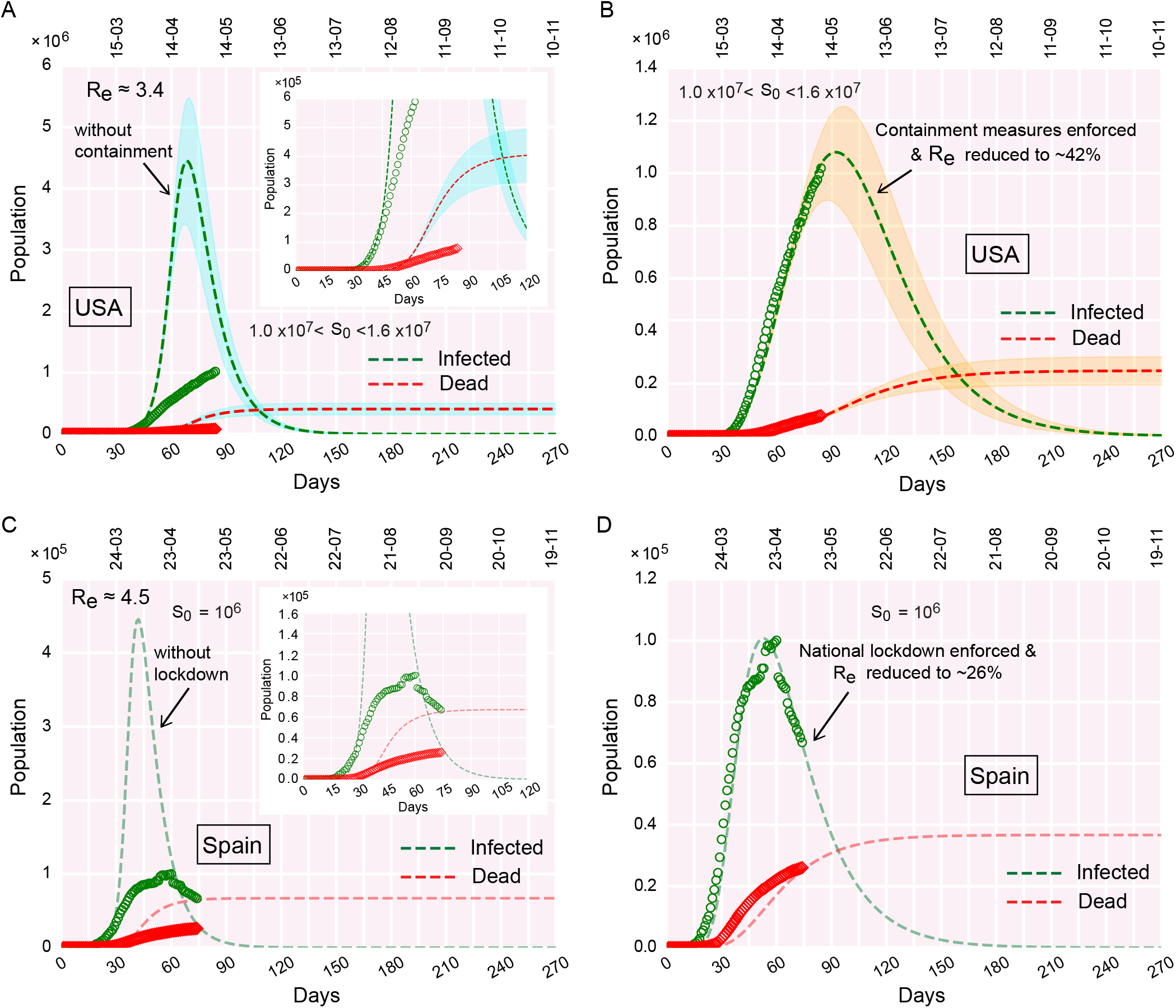
COVID-19 progression in the USA and Spain. (A-B) Time evolution of the population of infected and dead for the USA, (A) if containment measures were not undertaken, (B) due to the effect of containment measures implemented. Since for the USA, the infection peak is yet to occur, the estimation of the peak value is predictive, and thereby, sensitive to relevant parameter choices. Hence, the susceptible pool (*S*_0_) for the USA is varied, to obtain an average estimation of the peak value. The smeared color shades enveloping the dashed curves denote the variation in *S*_0_ in the context of the USA. (C-D) Time evolution of the population of infected and dead for Spain, (C) if containment measures were not undertaken, (D) due to the effect of containment measures implemented. The real data considered for fitting are from February 15, 2020, for the USA. For Spain, real data are considered from February 24.

**FIG. 9.**
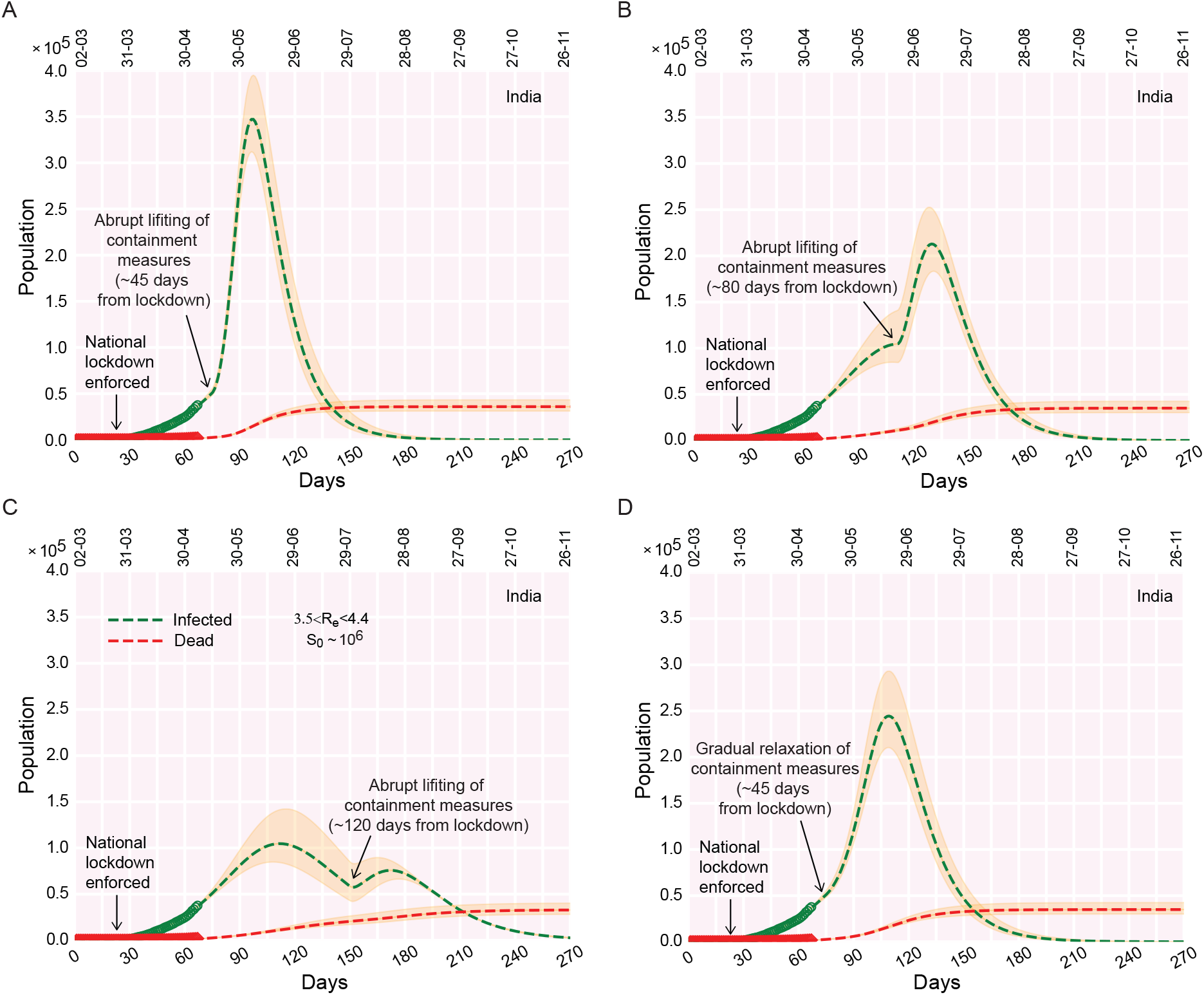
COVID-19 progression upon lifting the containment measures in India. (A-C) Time evolution of the population of infected, if the containment measures are ‘rapidly’ lifted ~ 45 days (A), ~ 80 days (B), ~ 120 days (C) from lockdown (e.g from the day lockdown was enforced). (D) Time evolution of the population of infected, if, ~ 45 days from lockdown, the containment measures are gradually relaxed/phased out within a time window of ~ 30 days. The real data considered for fitting are from March 2, 2020.

**FIG. 10.**
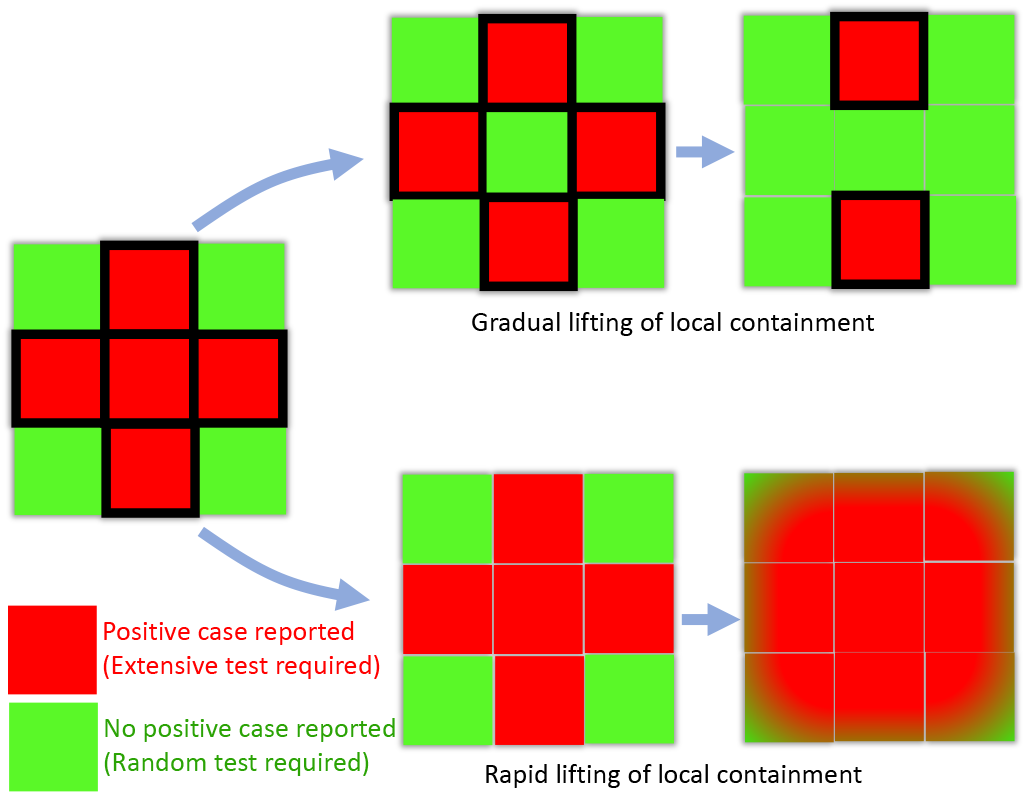
Representative schematic illustrating the aftermath of slowly relaxing (phasing out) of the local containment measures vs rapid lifting of the same. The boxes bordered in black depict the containment zones. Rapid lifting of the local containment paves the way for inter-mingling between regions with no positive cases (green) and regions with positive cases found lately (red). This, in turn, results in rapid transmission of the disease across zones (all zones becoming red), rendering the purpose of preceding lockdown futile. On the contrary, initially, the partial lifting of local containment only in the green zones bars the import of transmissions from red zones. When a red zone becomes green, the local containment can be lifted from that region. Due to the effect of this gradual lifting, the red zones diminish over time, with ‘greens’ taking over the ‘reds’.

### India without lockdown: What could have happened?

The first COVID-19 positive human host was reported in India on 30th January 2020. The exponential growth of the number of infections, from 30th January onward, reached a number 657 on 25th March 2020, the day on which India imposed a nation-wide lockdown (Fig. S1A).

Using the SIRD model, we first explored what could have happened, if the containment measures had not been undertaken. As mentioned earlier, we chose the factor ~ 10^−3^ (obtained in case of Germany and few other countries by dividing the cumulative population at infection peak by the actual population of the country) and multiplied it the Indian population of ~ 10^9^ to estimate the lower bound of the susceptible population (*S*_0_). It turns out that it would be a ‘good’ estimation to have a ‘working’ *S*_0_ in the range ~ 10^6^. With susceptible population *S*_0_ varied in the range ~ 1-3 million (for fixed 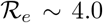), the peak of the infection occurs in the first half of May (Fig. 2A). As expected, the peak height (infected population at the peak) increases with increasing *S*_0_. For an initial susceptible pool of *S*_0_ ~ 10^6^, the peak reaches a height of 0.4 million, whereas the peak jumps to ~ 1.2 million for *S*_0_ ~ 3 million (Fig. 2A). The total death toll is estimated to reach about 30,000-100,000 for *S*_0_ in range ~ 1-3 million, during July - August, 2020 (Fig. 2A).

Next, we introduced the effect of containment-measures in the infection rate 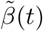 (Eq. 5-6). Numerical analysis is carried out to investigate whether the progression of the outbreak is mitigated after the lockdown is imposed.

### Effect of containment measures: How well is India doing?

#### Straightening of the growth curve

From the real data, it appears that the infection rate begins to reduce, 15-20 days after the national lockdown is implemented (Fig. 2A, inset). We further observed that the growth curve for the infected population displays a straightening feature during the lockdown time frame. This is expected to be observed if containment measures are initiated; the unhindered exponential growth before the lockdown slows down due to the effect of containment measures during the lockdown. While slowing down and deviating from the exponential trajectory, the infection growth curve (time progression of the infected population size) acquires a distinctive straightening feature until the very recent surge (Fig. 2A, inset).

#### Dwarfing the infection peak

Next, adding the lockdown effects into the picture, we fit the theoretically obtained infection growth curve with the real data. The best fit with the current set of parameters demonstrates that, due to the effect of the present lockdown, the infection peak dwarfs down to about 0.10 million from about 0.40 million in ‘without lockdown’ scenario (dashed curve, Fig. 2B and inset). The infection peak is projected to reach a peak at the end of June, tentatively (Fig. 2B, 2B, inset). The estimated death toll also reduces substantially compared to the earlier scenario without containment measures.

However, the model also shows that the situation can be improved further. The infection growth curve can be dwarfed down further if the lockdown is extended and reinforced stringently in COVID-19 prone zones. In that case, the infection growth curve noticeably flattens with the infection peak reduced further.

As mentioned earlier, *S*_0_ is a very crucial parameter in governing the position and the height of the infection peak. In the following, we summarize how the variations in the size of the susceptible population *S*_0_ influence the infection growth curve.

#### Variability in the susceptible population

Keeping 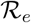 fixed at ~ 4.0, we varied the size of the total susceptible population within a range of 1-3 million (Fig. 2A-2B). The model analysis shows that the larger the size of the susceptible population, the higher the infection peak (Fig. 2). Moreover, for the larger size of the susceptible population, attainment of the infection peak is delayed with the infection peak shifted to a later time zone (Fig. 2B, 2B, inset). These characteristic features are consistent in both without and with lockdown scenarios.

Thus, it is evident that the key to containing the outspread lies in keeping *S*_0_ small. This is feasible only when interactions between a demographic region with the recent occurrence of infections and a region with no ‘latest’ instance of infection are strictly barred. Besides the ‘global’ lockdown (in a nation-wide sense), locally keeping an infected region isolated from other proximal unaffected regions may help to keep *S*_0_ in check.

The next question that crops up is what happens to the magnitude of effective reproduction number 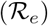 when containment measures are put in place. We discuss in the following, how the effective 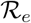 changes with time during the lockdown (Fig. 3).

#### Effective reproduction number 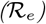 and lockdown

We start with 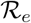 in range 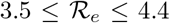 in the beginning. As the lockdown is implemented, less number of people interact. Therefore, the effective infection rate 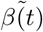 starts decreasing over time. How much the reduction would be for 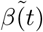 in longer time regime, is determined by the factor ζ in Eq. 5. The reduced 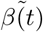 settles at a value ζ*β* due to the containment effects. Thus, if the re-covery rate *γ* is fixed, the 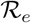 will diminish and reach a value 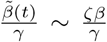. The decrease in 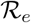 due to the effect of lockdown is evident in Fig. 3 where the effective 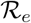 reduces to ~ 40 % of its initial value (before lockdown). The smeared color shades enveloping the dashed lines label the variations in 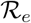 within the mentioned range. As expected, the higher the value of 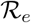, the taller the infection peak. This feature is consistent both in presence and absence of lockdown (Fig. 3, 3, inset).

Next, we investigate, whether the value of 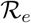, extracted from the best fit with real data, is unique (of course with marginal variation) or the variation is non-marginal.

#### What would be the scenario if 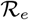 is large?

Instead of fixing 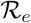 in the beginning, we varied the rates of infection (*β*), recovery (*γ*), and deaths (*δ*) without any restriction on the resulting value of 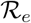. We aimed to verify whether the best fit of real data with the theoretical curves (infection, recovery, and death) can be obtained for a set of (*β*, *γ*, *δ*), other than the already chosen values in Fig. 2-3, with no apparent constraint on the values of 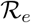.

We find that, for a fixed size of the susceptible population of about 0.8-1.2 million, the real data can still be fitted with the theoretical curves, even if the 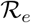 is large (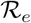 ~ 7.4, Fig. 4). Similar to Fig. 2-3, the active infection cases deviate from the theoretical infection curve without lockdown, as the enforced lockdown effectively slows down the progression of infection (Fig. 4, 4, inset). Note here, that in comparison with Fig. 2-3, the location and height of the infection peaks change (both in the cases of without lockdown and with lockdown), as effective 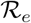 is increased ~ 2-fold (Fig. 4). Consistent with the definition of 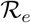, we observe that greater the value of 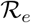, larger the size of the infected population (compare Fig. 2B, 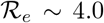; and Fig. 4, 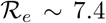). It is also noteworthy to mention that here the recovery rate *γ* = 0.0315 day^−1^ corresponds to about ~ 31 days compared to 12 days as discussed earlier. Prolonged infectiousness leads to the rise in the 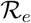 and consequently the total number of infected people.

From the above observations, we connote that the exactness of 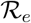 can be ascertained, when we have more data points in the time evolution of the infected, recovered, and dead population. The current model setup may not be able to precisely pinpoint the exact ‘real’ 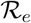.

#### How well the individual Indian states are doing?

In India, the first COVID-19 positive case was reported in Indian state Kerala on January 30, 2020, and now almost half of the active COVID-19 positive cases are from another Indian state Maharashtra. In this note, we explore the COVID-19 progression in these Indian states along with Delhi and West Bengal and compare the features of pandemic progression with each other (Fig. 5-6, S1B).

After the first case being detected in Kerala on January 30, the second and third cases were reported on Feb 2-3. After February 3, there was no new case detected in Kerala till March 7. The previous three cases were all recovered within February 20. The ‘second-wave’ of infections started from March 8. From March 8 onwards, there was a rapid upsurge of infections. However, about two weeks after the national lockdown is imposed, Kerala reached its infection peak. It is evident from Fig. 5A-5B that the downfall of the infection is rapid, as the infection curve moved past its peak. If the lockdown was not enforced, the infection was projected to occur around mid-May. But, Fig. 5B alludes that Kerala implemented the containment measures so well that the infection peak occurred early at a much lower height (Fig. 5A-5B). The model analysis further projects that due to the effect of lockdown, 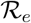 reduces to ~ 18 % of its initial value during the upsurge of infections before lockdown. The reduced value of 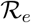 is << 1, which means Kerala is on the way to become a COVID-19 free state soon if the trend continues.

In Maharashtra, the first case was detected on March 9. The total infected population is yet to attain its peak. The projected infection peak would occur around the end of May or early June if the present trend continues and containment measures remain enforced in places (Fig. 5C-5D).

Similarly, in Delhi and West Bengal, the infection growth curves are yet to attain their respective peaks (Fig. 6A-6D). The first cases in these states were reported on March 2 and March 17 respectively. The peaks are projected to be reached at the end of June and mid-July for Delhi and West Bengal respectively, if the enforced lockdown remains deployed and the current trend continues (Fig. 6B, 6D).

It is important to note that, in Indian states, Maharashtra, Delhi and West Bengal, the estimated 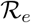 plummets down to value > 1.0, even after staying months under lockdown. Among the Indian states we analyzed, Kerala turns out to be the only exception where the effective 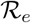 reduces to a value << 1, meaning that further ‘out-of-bound’ spreading is unlikely to occur there if the current trend is followed.

#### Progression of COVID-19: Where does India stand compared to other countries?

It is evident from Fig. 7A-7D and S1A that both Germany and South Korea have moved past the infection peak. The infected population is decreasing day by day in those countries. The best fit with real data is obtained for the initial 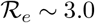 and ~ 4.5 for Germany and South Korea respectively. However, as the containment measures were undertaken in those countries, the effective transmission (or 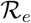) reduced to ~ 20-25 % from the initial values for the respective countries (Fig. 7B,7D). This observation, suggests that the counter-measures to fight the pandemic (e.g containment measures, social distancing, quarantining, testing, etc.), undertaken in these countries, were reasonably successful in repressing the outspread. Moreover, the reduction of 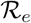 to 20-25 % of its initial values, rescales the 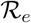 for the respective countries to a value < 1 which alludes that new infections are declining and any more ‘out-of-bound’ infection growth is unlikely to occur if the current trend is followed.

We analyzed COVID-19 progression data for two more countries: USA and Spain (Fig. 8A-8D). The USA is approaching the infection peak and will reach its peak shortly if the current trend continues (Fig. 8B). However, contrary to the USA, Spain has already passed the infection peak and the infected population is decreasing gradually (Fig. 8D). Spain imposed a nation-wide lockdown on March 14. Model analysis (fitting parameter optimization) suggests that, due to the effect of lockdown, 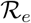 for Spain reduced to 25 % of its initial value. But in the USA, the reduction in 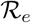 is only ~ 42 % implying that the implementation of local containment was not that stringent.

Contrary to the infection curves for Germany, South Korea, Spain (Fig. 7-8), India is yet to reach the infection peak (the USA is about to reach the peak.). The following remarks briefly summarize where India stands compared to these countries.

1. The effective containment during the present lockdown in India indicates that the infected population might reach its peak at the end of June (Fig. 2) whereas Germany, South Korea, and Spain have already moved past the peak and daily new infections are decreasing (Fig. 7-8).
2. Since India has a large population, the infection is expected to stay for a longer duration. Germany, South Korea, and Spain might have the advantage of a smaller population of the susceptible. We allude that the higher the actual population of a country, the higher would be the effective size of the susceptible pool for that country while making the previous statement. The key is to contain the infections in small zones and prevent transmission between infectious and non-infectious zones.
3. The growth of the infected population in Germany, South Korea, and Spain were greater than that experienced in India which gave India an additional advantage of ‘buying precious time’. Slow growth rate alludes to a smaller peak value at the zenith of the infection. However, as mentioned earlier, the height of the peak is subjected to the effective size of the initial susceptible pool (*S*_0_).

## DISCUSSION

For better clarity and wider accessibility to general readers, we discuss and summarize the important observations from our study in Q&A format in the following:

### How accurate are the model predictions?

The SIRD model is a drastically simplified approach to thoroughly understand the dynamics of COVID-19 progression. From the available data, it is now clear that a susceptible person goes through a latent period of 2-3 days after coming in contact with an infected individual. Subsequently, the person remains infectious for several days (~5 days). The infectious individual may or may not develop symptoms. The current model does not incorporate any of these details and hence fitting is imperfect. Moreover, data used to fit with the model also vary between different locations leading to uncertain predictions. A compartmental model with multiple species may be useful to study the dynamics of the subpopulation [13].

#### How sensitive are the model predictions to parameter variations?

We investigated the sensitivity of the model to parameter variations, focusing in particular on the parameters that change the rates of infection, recovery (*β*, *γ*) and most importantly the effective reproduction number or replacement number 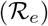 within a feasible range to see the effect the model prediction. The lockdown stringency, characterized by the time-dependent *β*, ζ, was varied to get an estimate of the infected population. Corresponding data are shown in each figure by the shaded envelope around the mean curves. The model appears to be sensitive to the variation in the value of ζ and *S*_0_ when compared with the real data. Increasing (decreasing) the value *S*_0_, ζ, and *R*_0_ rapidly increases (decreases) the population of total infection and death mostly around the peak and alters the position of the peak infection early. These parameters can be decreased by enforcing local containment and social-distancing measures.

### How universal the model predictions are for different countries?

To investigate the universality in the COVID-19 outspread across different countries, we looked into iterative time lag maps for the cumulative confirmed infected (C = I+R+D), recovered (R), and dead (D) population [6]. Using the iterative maps, we try to extract the correlation between a population on the day *n* and day *n* + 1. From the recurrence plots (population count on *n^th^* day vs population count on (*n* + 1)*^th^* day) in Fig. S2, we observe that the real data for all the cases follow the same power law of the following kind: *f* (*x*) = *ax^b^*. The factor and exponent *a* and *b* are similar for all the countries considered in the plots. This finding indicates that there exists an underlying universality in the outspread of the pandemic across various countries.

### How effective are the quarantine measures? Can quarantining single-handedly contain the transmission?

A common perception of flu and other infectious diseases is that an infected individual spreads infection when symptoms appear. In the case of seasonal flu, infection mostly occurs when a person has symptoms [14]. However, as we understand from the literature survey, an individual with COVID-19 would be contagious before developing symptoms. The incubation period for COVID-19 is ~ 5 days, and maximum infectiousness appears to be 2-3 days before the symptoms appear. Thus infection spread by an individual is maximum before he/she becomes sick [10, 15]. Due to limitations of the testing procedure, diagnosis takes about 5 days after symptoms are visible, i.e., 10 days from the day of infection. Clearly, on the average, an infected individual is beyond the peak of maximum infectiousness after this time. Thus, a reduced rate of infection demands early diagnosis and isolation of positive patients. This means that a COVID-19 patient needs to be identified in the pre-symptomatic stage as evidence suggests the infectiousness of the patient before developing symptoms which is extremely challenging (effectively, RT-PCR needs to be carried out for every individual who might have come in contact with the patient). The epidemic becomes even more complex due to a majority of the infected individual who develops mild or no symptoms [15, 16]. Therefore, even with isolating/quarantining, all the infected COVID-19 would not be eliminated for two reasons: a) normally an individual would be tested after symptoms appear which is when he/she has passed the peak of the contagiousness, b) asymptomatically infected person, in general, are not tested but he/she is also contagious like the symptomatic individual.

### Gradual relaxation of the containment versus extended lockdown?

Relaxing the containment: We have investigated the effect of relaxing the containment measures at 3 different time points for India (Fig. 9A-9D). We find that if the containment is relaxed before the peak infection is reached at the end of June, the infection would rise rapidly to a great extent (Fig. 9A). The peak height reduces, if the containment measures are relaxed, when the infection is close to the peak, a time point around the 3rd week of June (Fig. 9B). However, if the relaxation occurs a month after the peak infection, a second peak arrives which is lower than the first infection-peak (Fig. 9C). A third possibility is to gradually relax the containment measures after May 17. The model shows that in this case the original peak does not shift its position but becomes two-fold higher than before (Fig. 9D). A gradual relaxation could be carried out in steps: (a) First, identify all the sensitive (red) and safe (green) zones having positive and no cases respectively. Smaller the size of such zones, easier they can be managed by the administration, and necessary supplies can be arranged. It is important to seal the boundary of the red zones. (b) Test for new cases carrying symptoms and randomly test a few having no symptoms. (c) Dissolve the boundary between red and neighboring green zones once the red zone does not report a case for two weeks. This process will increase the size of the green zone where more and more people can communicate and business can restart. Successively extending the relaxation from the local neighborhood to the cities, districts, states, the containment measures can be relaxed across the country. Nevertheless, social distancing is mandatory even after the containment is officially lifted as there might be many undetected cases that can trigger the spread of the disease again. A schematic diagram in Fig. 10 summarizes the above-mentioned steps of relaxation and the consequential aftermaths, pictorially. In a nutshell, ‘too-early’ lifting of containment measures, long before the infection reached its peak, makes the purpose of lockdown ‘null-and-void’. The reduced infection rate 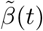 again starts increasing yielding a larger 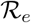 promoting the outspread.

Our model projects that, with containment measures in effect, the number of active infection would be about 80,000 - 140,000 at the peak which is expected to appear sometime after mid-June (Fig. 3). Note that, due to containment the number of susceptible populations decreases drastically to only several million while the total population of India is about 1.35 billion. If the lockdown is suddenly relaxed locally after May 17, the peak infection would rise sharply to 0.3 - 0.4 million (Fig. 9A). It is imperative to mention that, this number is estimated without altering the susceptible population which is about 10^−3^ times the actual population observed for many large countries. The remaining population considered to be shielded from the infection due to containment and demographic segregation. In the event of unrestricted mixing of the population of the whole country, the peak infection might see a 10^3^ fold rise which would be challenging for any health care system to deal with. Thus, social distancing measures must remain in place unless the infectious population is drastically contained.

### What is causing the local resurgence of positive cases even after the 7 weeks of extended lockdown?

Although the nation is under lockdown, it is observed that the number of positive cases is still growing at large. A distinct feature of this growth is the local resurgence of infections. As gleaned from various news reports, even after several days with a few new cases, suddenly, there had been jumps in the COVID-19 positive cases in quite a few places. In other words, ‘lull’ ‘green’ zones are, all of a sudden, turning into ‘red’ zones. We discuss a few plausible factors behind the resurgence: (a) Cross-country reverse migration: Due to the lockdown, a large population of migrant workers reeling at the bottom of the economic barrel got stranded in different places without much subsistence. These people started returning to their homes taking desperate measures. During this migration, human-to-human transmission of COVID-19 might have occurred to a great extent due to a lack of social distancing adding fuel to the ‘resurgence’ of infection. (b) Lack of ‘test, trace and contain’: Interestingly, an important aspect of COVID-19 is the number of patients who do not develop any symptoms (Fig. 1B). In India, primarily the testing capacity was devoted to the persons showing typical symptoms of COVID-19. The asymptomatic pool largely remained unnoticed at the initial stages of infection outgrowth which probably contributed to the resurgence of infections. Moreover, it is not sufficient to only isolate the positive cases but to trace all those people who came in contact with the individual tested positive and find the source of infection. This is known as ‘contact tracing’. If the source of the infection is not traceable, this could indicate an insufficient testing or asymptomatically positive source. Extensive use of the app-based modern technology may become useful to trace contacts, however, often at the cost of privacy. In India, where a large portion of the population has no ‘digital footprint’, contact tracing becomes even harder. South Korea flattened the infection curve with extensive testing and other mentioned measures. In India, a similar endeavor of a magnitude proportional to its humongous population seems extremely challenging. With the limited capacity and huge population, randomized testing, at least in the infectious neighborhood, is an immediate solution to detect and isolate the asymptomatic individuals.

In both the above-mentioned scenarios, the majority of the infection spreading is likely to be spearheaded by the asymptomatic human hosts who remain undetected due to a lack of randomized testing and come in social contact with others. This means that they would be infecting healthy people unknowingly. According to the WHO and the Indian Council of Medical Research (ICMR), as much as 80% of the infected individual can be asymptomatic. Thus, all the symptomatic cases reported so far contribute to only about 20% of the total infection. Going by the reported number of cumulative infections 61,356 as on May 9, almost all of which are symptomatic, this would correspond to about 245,424 people who also had the virus but did not show any symptoms. Together, about 306,780 people have actually been infected so far in India carrying symptoms or no symptoms. Therefore, the number of people in the country who are still susceptible to the infection is still in the order of billion. One can realize that, with so many active infections, extensive mixing of the countrywide population soon after the lockdown is over (after 17 May) would cause a huge surge in the total number of infections which is nearly impossible to manage by any health care system. In order to estimate the asymptomatic population from the model, we rewrite the equations as follows:

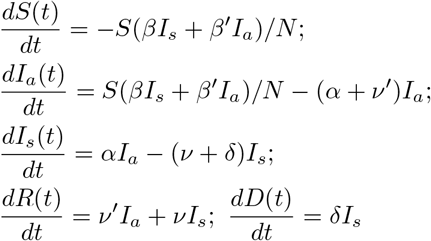

Here the total infectious population is segregated into two compartments: (a) symptomatic *I_s_* and (b) asymptomatic or mildly symptomatic *I_a_* population. A susceptible person can be infected upon contact with a symptomatic or asymptomatic individual with rates *β*, *β′* respectively. The infected individual can remain asymptomatic or mildly symptomatic and transit into a symptomatic state with rate *α*. The asymptomatic and symptomatic persons can recover at rates *ν*′ and *ν* respectively. For a symptomatic individual, death occurs with rate *δ*. For simplicity, we assumed no death for asymptomatic population. We find that with an *S*_0_ in range ~ 4-6 million and 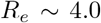, new model data (Fig. S3) matches with the previously plotted population of symptomatic infection (see Fig. 2). The asymptomatic infection peak appears to be about 4 fold larger than the symptomatic infection peak. Thus the current lockdown can only be relaxed in the presence of extensive testing of symptomatic and asymptomatic population and contact tracing.

It is noteworthy to mention that the total number of cases reported in all over India as well as in various Indian states are negligible compared to the total population of the country and states respectively. Besides, the severity of the infection with symptoms is relatively less in India than in the USA and other large European countries. Whether it is due to the effect of hot and humid weather of India or other meteorological parameters such as high UV index, future research would be able to evaluate.

## Data Availability

The real data for analysis in India's context is collected from the repository with an interactive interface hosted at https://www.covid19india.org. The data for other countries are taken from the repository with an interactive interface hosted at https://www.worldometers.info/coronavirus.

https://www.covid19india.org

https://www.worldometers.info/coronavirus

## ACKNOWLEDGMENTS

We thank Santanu Bhattacharya, Heiko Rieger, Soumitra Sengupta, Jayanta Kumar Bhattacharjee, Deb Shankar Ray, Sankar Prasad Bhattacharyya, Parongama Sen for exciting discussions and valuable suggestions. S.C.* and A.S. were supported by a fellowship from the University Grants Commission (UGC), India. S.C.^‡^ thanks Indian Association for the Cultivation of Science, Kolkata for financial support. M.K. was supported by a fellowship from CSIR, India. R.P. thanks IACS for support and Grant No. EMR/2017/001346 of SERB, DST, India for the computational facility.

---

**FIG S1.**
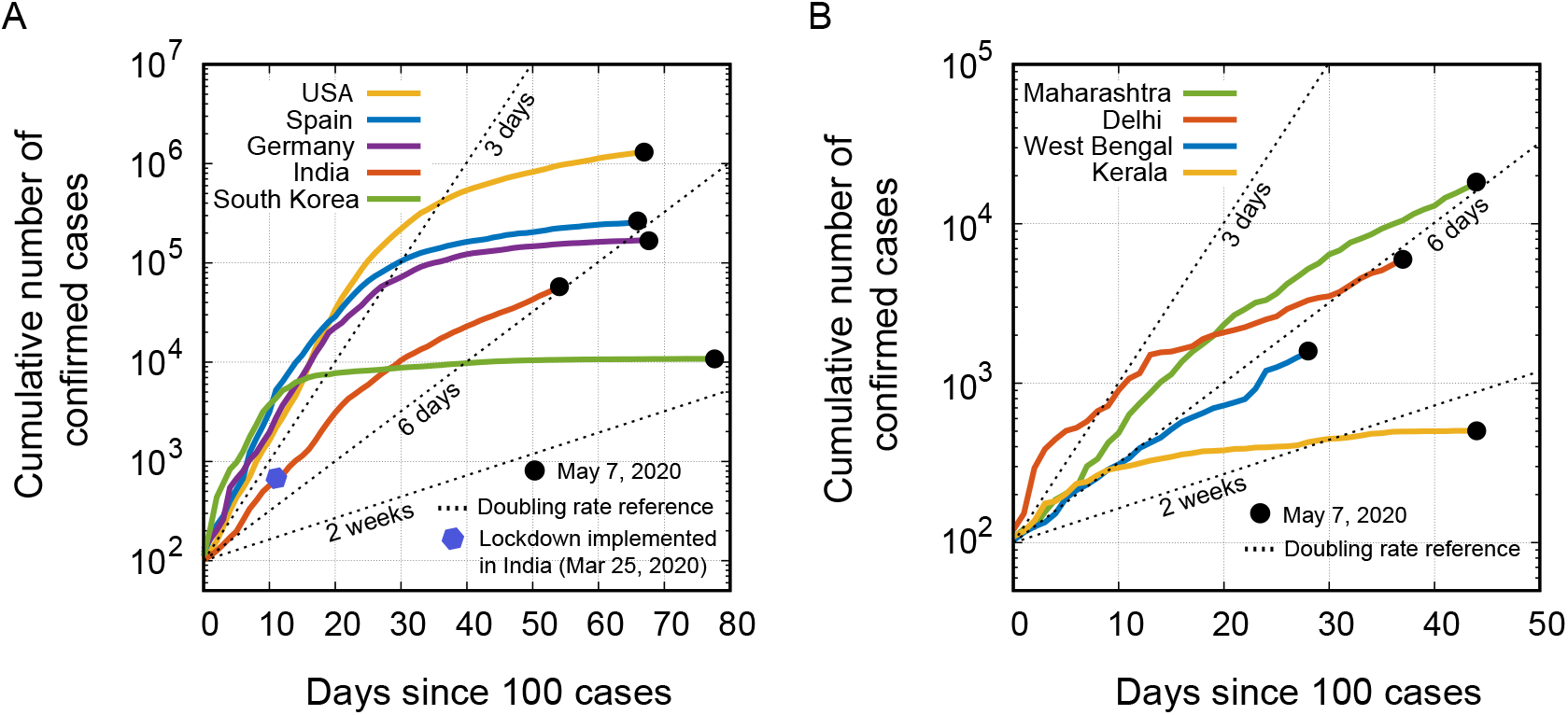
Time progression of the cumulative number of confirmed COVID-19 positive cases from the days since 100 cases for (A) different countries (B) different Indian states, plotted on a semi-log scale.

**FIG S2.**
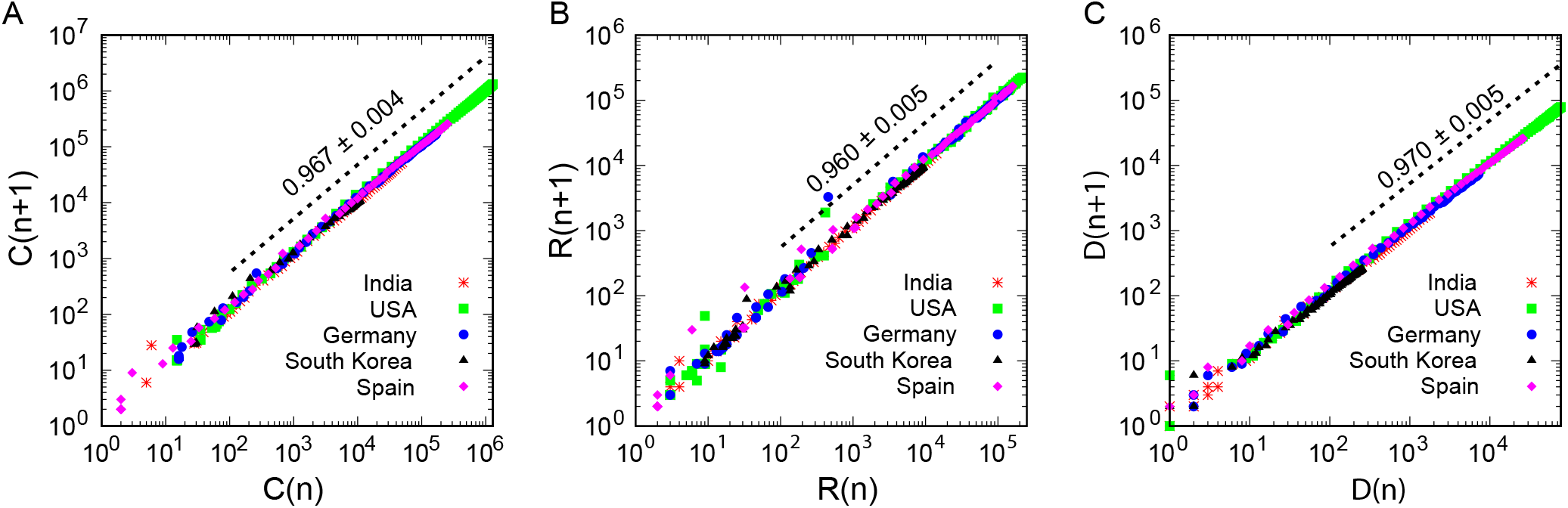
The recurrence correlation between the population on day *n* and day *n* + 1 plotted on a log-log scale. (A) *C*(*n*) stands for the number of cumulative confirmed infective cases on day *n*. (B) *R*(*n*) stands for total recovered population on day *n*. (C) *D*(*n*) stands for total deaths on day *n*. The population count on *n^th^* day vs population count on (*n* + 1)*^th^* day plots for several countries, exhibit same power law of the following kind: *f* (*x*) = *ax^b^*.

**FIG S3.**
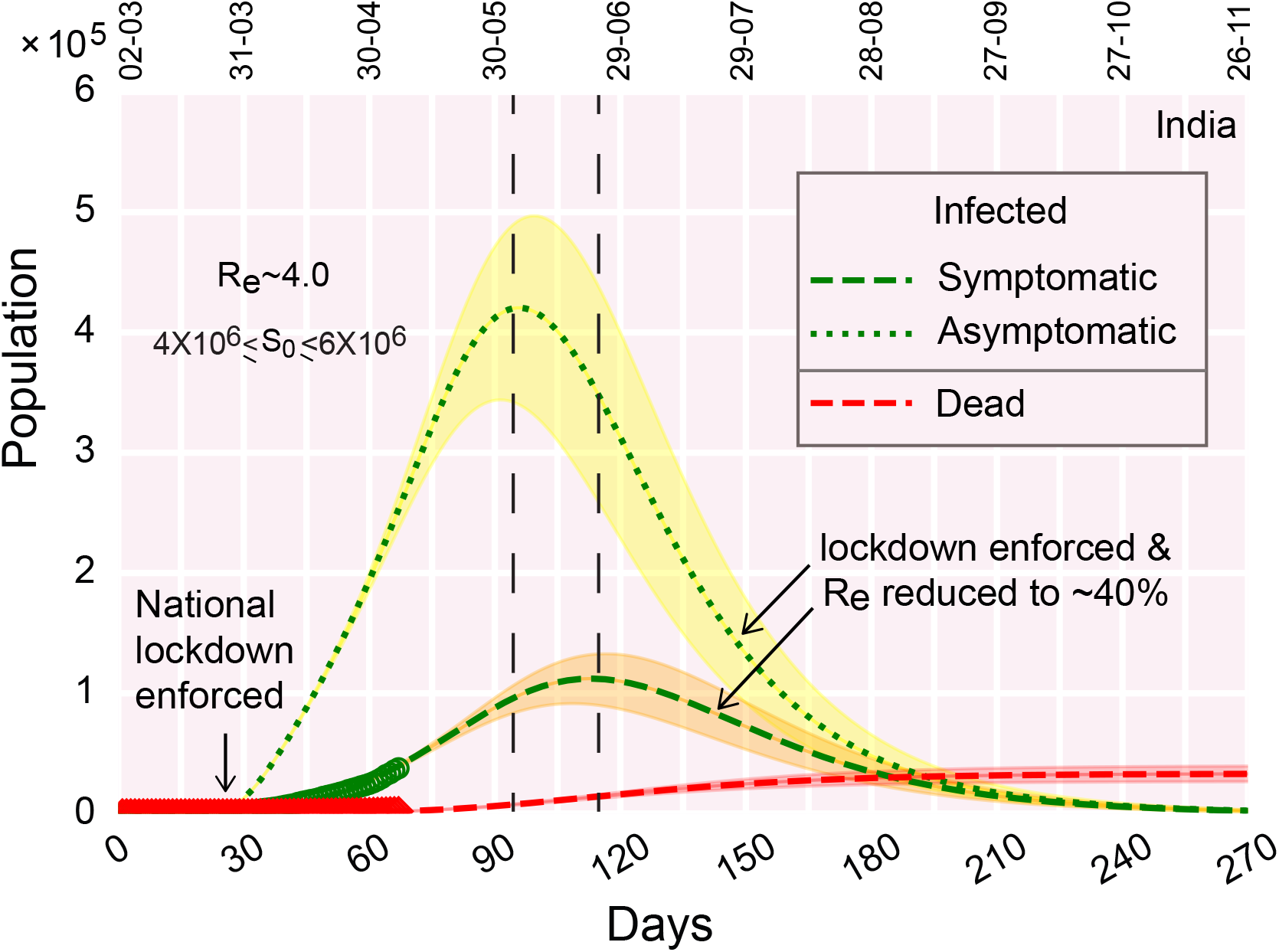
Infection growth curve in Indian context when the outspread is transmitted through both symptomatic and asymptomatic subpopulation. Relevant rate parameters (day^−1^): *β* = 0.18, *β′* = 0.341, *ν* = 0.045, *ν′* = 0.085, *δ* = 0.0031, *α* = 0.015; lockdown related parameters: *τ* = 30 days, *T* = 7 days, ζ = 0.42.

**TABLE S1.** List of parameters chosen for the best fit with real data in Indian context.

| Abbreviation | Meaning | Value |
| --- | --- | --- |
| **Parameters for India** |  |  |
| $\beta$ | rate of infection | $0.2945 \text{ day}^{-1}$ |
| $\gamma$ | rate of recovery | $0.073 \text{ day}^{-1}$ |
| $\delta$ | rate of death | $0.0028 \text{ day}^{-1}$ |
| $\zeta$ | infection/interaction parameter | 0.435 |
| $\tau$ | The day from which the containment measures are implemented counted from day 0 | 30 days |
| $T$ | delay in number of days before the effect of containment measures becomes prominent | 9 days |
| $S_0$ | initial susceptible population | $1.0\text{-}3.0 \times 10^6$ |

**TABLE S2.** List of parameters chosen for the best fit with real data in Indian states Kerala, Maharashtra, West Bengal and Indian capital territory Delhi.

| Abbreviation | Meaning | Value |
| --- | --- | --- |
| Parameters for<br>Indian state **Kerala** |  |  |
| $\beta$ | rate of infection | $0.26 \text{ day}^{-1}$ |
| $\gamma$ | rate of recovery | $0.092 \text{ day}^{-1}$ |
| $\delta$ | rate of death | $0.004 \text{ day}^{-1}$ |
| $\zeta$ | infection/interaction parameter | 0.18 |
| $\tau$ | The day from which<br>the containment measures<br>are implemented<br>counted from day 0 | 22 days |
| $T$ | delay in number of days before<br>the effect of containment<br>measures becomes prominent | 4 days |
| $S_0$ | initial susceptible population | $10^4$ |
| Parameters for<br>Indian state **Maharashtra** |  |  |
| $\beta$ | rate of infection | $0.27 \text{ day}^{-1}$ |
| $\gamma$ | rate of recovery | $0.0576 \text{ day}^{-1}$ |
| $\delta$ | rate of death | $0.0038 \text{ day}^{-1}$ |
| $\zeta$ | infection/interaction parameter | 0.55 |
| $\tau$ | The day from which<br>the containment measures<br>are implemented<br>counted from day 0 | 20 days |
| $T$ | delay in number of days before<br>the effect of containment<br>measures becomes prominent | 15 days |
| $S_0$ | initial susceptible population | $0.8-1.6 \times 10^5$ |
| Parameters for<br>Indian state **West Bengal** |  |  |
| $\beta$ | rate of infection | $0.314 \text{ day}^{-1}$ |
| $\gamma$ | rate of recovery | $0.086 \text{ day}^{-1}$ |
| $\delta$ | rate of death | $0.008 \text{ day}^{-1}$ |
| $\zeta$ | infection/interaction parameter | 0.452 |
| $\tau$ | The day from which<br>the containment measures<br>are implemented<br>counted from day 0 | 14 days |
| $T$ | delay in number of days before<br>the effect of containment<br>measures becomes prominent | 13 days |
| $S_0$ | initial susceptible population | $0.8-1.6 \times 10^5$ |
| Parameters for<br>India's capital territory **Delhi** |  |  |
| $\beta$ | rate of infection | $0.24 \text{ day}^{-1}$ |
| $\gamma$ | rate of recovery | $0.06 \text{ day}^{-1}$ |
| $\delta$ | rate of death | $0.004 \text{ day}^{-1}$ |
| $\zeta$ | infection/interaction parameter | 0.54 |
| $\tau$ | The day from which<br>the containment measures<br>are implemented<br>counted from day 0 | 24 days |
| $T$ | delay in number of days before<br>the effect of containment<br>measures becomes prominent | 13 days |
| $S_0$ | initial susceptible population | $0.8-1.6 \times 10^5$ |

**TABLE S3.**
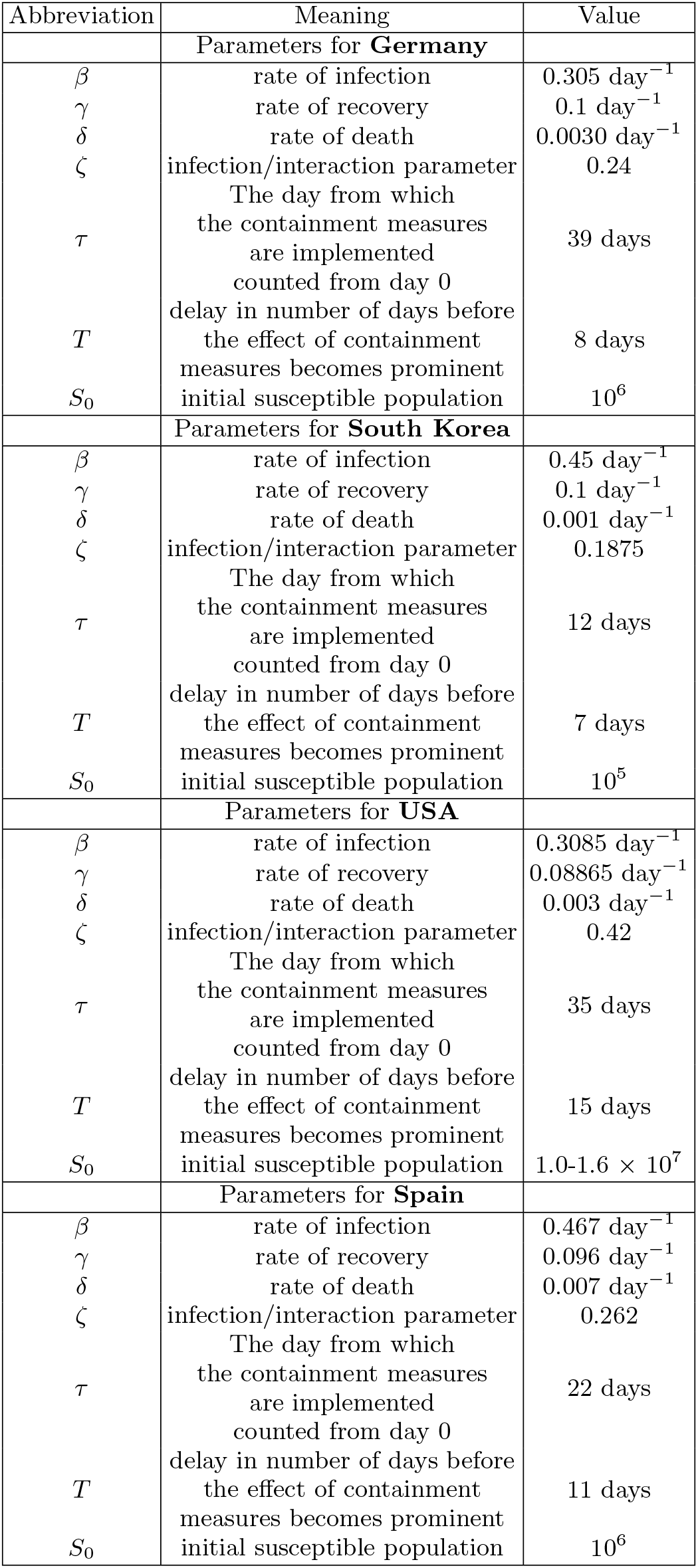
List of parameters chosen for countries Germany, South Korea, USA and Spain.

| Abbreviation | Meaning | Value |
| --- | --- | --- |
| **Parameters for Germany** |  |  |
| $\beta$ | rate of infection | $0.305 \text{ day}^{-1}$ |
| $\gamma$ | rate of recovery | $0.1 \text{ day}^{-1}$ |
| $\delta$ | rate of death | $0.0030 \text{ day}^{-1}$ |
| $\zeta$ | infection/interaction parameter | 0.24 |
| $\tau$ | The day from which the containment measures are implemented counted from day 0 | 39 days |
| $T$ | delay in number of days before the effect of containment measures becomes prominent | 8 days |
| $S_0$ | initial susceptible population | $10^6$ |
| **Parameters for South Korea** |  |  |
| $\beta$ | rate of infection | $0.45 \text{ day}^{-1}$ |
| $\gamma$ | rate of recovery | $0.1 \text{ day}^{-1}$ |
| $\delta$ | rate of death | $0.001 \text{ day}^{-1}$ |
| $\zeta$ | infection/interaction parameter | 0.1875 |
| $\tau$ | The day from which the containment measures are implemented counted from day 0 | 12 days |
| $T$ | delay in number of days before the effect of containment measures becomes prominent | 7 days |
| $S_0$ | initial susceptible population | $10^5$ |
| **Parameters for USA** |  |  |
| $\beta$ | rate of infection | $0.3085 \text{ day}^{-1}$ |
| $\gamma$ | rate of recovery | $0.08865 \text{ day}^{-1}$ |
| $\delta$ | rate of death | $0.003 \text{ day}^{-1}$ |
| $\zeta$ | infection/interaction parameter | 0.42 |
| $\tau$ | The day from which the containment measures are implemented counted from day 0 | 35 days |
| $T$ | delay in number of days before the effect of containment measures becomes prominent | 15 days |
| $S_0$ | initial susceptible population | $1.0\text{-}1.6 \times 10^7$ |
| **Parameters for Spain** |  |  |
| $\beta$ | rate of infection | $0.467 \text{ day}^{-1}$ |
| $\gamma$ | rate of recovery | $0.096 \text{ day}^{-1}$ |
| $\delta$ | rate of death | $0.007 \text{ day}^{-1}$ |
| $\zeta$ | infection/interaction parameter | 0.262 |
| $\tau$ | The day from which the containment measures are implemented counted from day 0 | 22 days |
| $T$ | delay in number of days before the effect of containment measures becomes prominent | 11 days |
| $S_0$ | initial susceptible population | $10^6$ |

